# Increasing but inadequate intention to receive Covid-19 vaccination over the first 50 days of impact of the more infectious variant and roll-out of vaccination in UK: indicators for public health messaging

**DOI:** 10.1101/2021.01.30.21250083

**Authors:** Patrik Bachtiger, Alexander Adamson, William A Maclean, Jennifer K Quint, Nicholas S Peters

## Abstract

**Objectives:** To inform critical public health messaging by determining how changes in Covid-19 vaccine hesitancy, attitudes to the priorities for administration, the emergence of new variants and availability of vaccines may affect the trajectory and achievement of herd immunity.

**Methods:** >9,000 respondents in an ongoing cross-sectional participatory longitudinal epidemiology study (LoC-19, n=18,581) completed a questionnaire within their personal electronic health record in the week reporting first effective Covid-19 vaccines, and then again after widespread publicity of the increased transmissibility of a new variant (November 13th and December 31st 2020 respectively). Questions covered willingness to receive Covid-19 vaccination and attitudes to prioritisation. Descriptive statistics, unadjusted and adjusted odds ratios (ORs) and natural language processing of free-text responses are reported, and how changes over the first 50 days of both vaccination roll-out and new-variant impact modelling of anticipated transmission rates and the likelihood and time to herd immunity.

**Findings:** Compared with the week reporting the first efficacious vaccine there was a 15% increase in acceptance of Covid-19 vaccination, attributable in one third to the impact of the new variant, with 75% of respondents “shielding” – staying at home and not leaving unless essential – regardless of health status or tier rules. 12.5% of respondents plan to change their behaviour two weeks after completing vaccination compared with 45% intending to do so only when cases have reduced to a low level. Despite the increase from 71% to 86% over this critical 50-day period, modelling of planned uptake of vaccination remains below that required for rapid effective herd immunity – now estimated to be 90 percent in the presence of a new variant escalating R_0_ to levels requiring further lockdowns. To inform the public messaging essential therefore to improve uptake, age and female gender were, respectively, strongly positively and negatively associated with wanting a vaccine. 22.7% disagreed with the prioritisation list, though 70.3% were against being able to expedite vaccination through payment. Teachers (988, 12.6%) and Black, Asian and Minority Ethnic (BAME) (837, 10.7%) groups were most cited by respondents for prioritisation.

**Interpretation:** In this sample, the growing impact of personal choice among the increasingly informed public highlights a decrease in Covid-19 vaccine hesitancy over time, with news of a new variant motivating increased willingness for vaccination but at levels below what may be required for effective herd immunity. We identify public preferences for next-in-line priorities, headed by teachers and BAME groups, consideration of which will help build trust and community engagement critical for maximising compliance with not only the vaccination programme but also all other public health measures.

## INTRODUCTION

With a higher burden of Covid-19 infection and a greater threat than ever of health services worldwide being overwhelmed, vaccination programs provide the route to ending the crisis phase of the pandemic by creating herd immunity.^1^ But achieving herd immunity and its rate of attainment are dependent on intended uptake of vaccination which not only remains unknown but is likely to have changed with both materialisation of vaccination programmes and the coincident emergence of substantially more infectious variants. Messaging around both of these widely publicised events individually, and in combination with pre-existing biases and attitudes to vaccination, needs to be effectively directed to maximise vaccine uptake as the only immediately modifiable and critical variable in the drive for herd immunity.^2,3^

Given the growing impact of personal choice among the increasingly informed public^4^, and extensive media coverage of both vaccine development and the increased infectivity of new variants, we aimed to characterise attitudes to the COVID-19 vaccine from the responses of more than 9,000 patients completing a questionnaire within the UK’s biggest personal health record in the week of approval of the first vaccination (13th November 2020). The questionnaire was repeated after the critical first 50 days of widespread publicity of both vaccine availability and roll-out, and the increased risk of infection and rapidly escalating case numbers from the new variant that led to the UK Government’s decision to impose the New-Year lockdown five days later.

Community engagement is more important than ever to maximise compliance with public health measures^5^ and requires acceptance of the current UK government prioritisation list for Covid-19 vaccination^6^, and for the remaining majority to not only accept vaccination but also the delay before they receive it. Public perception of the prioritisation and who should be next in line beyond the current list remains unknown but is an important consideration for informing health policy that builds public trust and compliance not only with the vaccination programme, but critically also with all other public health measures to reduce the impact of the pandemic.

Given the critical importance of maximising vaccine uptake as the vaccination programme gathers pace and new variants continue to emerge, we sought to determine how this changing environment changes Covid-19 vaccine hesitancy and attitudes to both vaccination, the priorities for administration and modelling of the trajectory and acquisition of herd immunity.

## METHODS

### Study participants

The Longitudinal Effects on Wellbeing of the Covid-19 Pandemic (LoC-19) study is an ongoing participatory epidemiology study with registrants (n = 18,581) receiving weekly questionnaires to complete within their personal electronic health record, the Care Information Exchange (CIE) of Imperial College Healthcare NHS Foundation Trust (ICHNT). The CIE is the NHS’ largest patient-facing electronic health record with UK-wide registrants (supplementary figure 1). Registrants were previously able to opt in to be LoC-19 participants by questionnaire completion between April 9th and June 5th 2020.

### Questionnaire Design and Timing

Applying recommendations for questionnaire design^7,8^, question items were developed by a collaboration of experts in qualitative research at Imperial College London, encompassing public health, respiratory epidemiology and digital health. Question items were externally peer-reviewed and tested on lay persons (n = 5) before being included in the final questionnaire.

Questionnaires were sent to participants on November 13th (week Pfizer BioNTech vaccine efficacy reported >90%) and December 31st 2020 (two weeks after first reports of new, more transmissible variant of Covid-19). The first questionnaire covered willingness to receive Covid-19 vaccination and attitudes to prioritisation, including free-text option for next-in-line prioritisation. The UK government’s Covid-19 vaccine prioritisation list was presented within the questionnaire. The second covered willingness to receive Covid-19 in the context of an emergent, highly transmissible variant, and plans for behaviour change after vaccination.

### Inclusion and Exclusion Criteria

Responses submitted later than four days from the time of the questionnaire launch were excluded.

### Data analysis

Descriptive statistics and unadjusted and adjusted odds ratios (ORs) were processed using R (version 3.6.2). and reported alongside machine learning analysis (natural language processing in Python version 3.7) of free-text responses. For the latter, tokenisation using SpaCy’s tokenizer was followed by rules-based labelling using RegEx and SpaCy’s token matcher, with each individual label verified by the authors. Inclusion was limited to labels (groups) with >200 responses.

## RESULTS

### First Questionnaire (November 13th, 2020)

Among 9122 respondents (49.4% response rate within time limit), 6521 (71.5%) would want Covid-19 vaccination, and 880 (9.6%) would refuse. Although 2068 (22.7%) disagreed with the government’s order of priority, 6416 (70.3%) were against being able to pay for vaccination (table 1). Excluding those uncertain (N = 1678), yearly increase in age (unadjusted-OR: 1.050 (95%CI:1.044-1.055), adjusted-OR: 1.045 (95%CI:1.039-1.050), and female gender (unadjusted- OR 0.415 (95%CI:0.356-0.482), adjusted-OR 0.540 (95%CI:0.461-0.632)) increased and decreased vaccine acceptance respectively, such that a yearly increase in age was associated with a 5% increase in likelihood of wanting vaccination. Overall, 64.2% of females would want vaccination and 12.4% would not, compared to 80.8% and 6.5% of males, respectively.

**Table 1.** Baseline demographics, vaccine hesitancy and attitudes to prioritisation list. First Questionnaire (November 13th, 2020).

|  |  |
| --- | --- |
| **Total N (%)** | 9122 (100) |
| **Age Mean (SD)** | 59.7 (13.6) |
| **Age category N (%)** |  |
| **18-29** | 173 (1.9) |
| **30-39** | 749 (8.2) |
| **40-49** | 1123 (12.3) |
| **50-59** | 2065 (22.6) |
| **60-69** | 2606 (28.6) |
| **70-79** | 2017 (22.1) |
| **80+** | 389 (4.3) |
| **Gender** |  |
| **Male** | 4176 (45.8) |
| **Female** | 4945 (54.2) |
| **Indeterminate** | <5 (0.0) |
| **Ethnicity** |  |
| **White background** | 6210 (68.1) |
| **Non-White background** | 1256 (13.8) |
| **Prefer not to say** | 86 (0.9) |
| **Missing\*** | 1570 (17.2) |
| **If a COVID-19 vaccine was available, would you want to have it?** |  |
| **Yes** | 6521 (71.5) |
| **No - because I believe I have health conditions that make it unsafe** | 257 (2.8) |
| **No - because I don't believe in vaccination** | 71 (0.8) |
| **No - because I think I have had COVID-19 infection (but not tested)** | 88 (1.0) |
| **No - because testing has confirmed I have had COVID-19 infection** | 68 (0.7) |
| **No - other** | 396 (4.3) |
| **Not sure** | 1678 (18.4) |
| **Missing** | 43 (0.5%) |
| **Do you think you should be allowed to pay to have a COVID-19 vaccine without delay?** |  |
| **Yes** | 2435 (26.7) |
| **No** | 6416 (70.3) |
| **Missing** | 271 (3.0) |
| **Do you agree that this order of priorities is correct?\*\*** |  |
| **Yes** | 6798 (74.5) |
| **No** | 2068 (22.7) |
| **Missing** | 256 (2.8) |
| **Are you concerned that the government list makes no direct reference to Black and Asian and Minority Ethnic groups?\*\*\*** |  |
| **Yes** | 2971 (32.6) |
| **No** | 5823 (63.8) |
| **Missing** | 328 (3.6) |

\*Ethnicity reported by participants in a previous LoC-19 questionnaire, not completed by 17.2% of participants of the 13th November 2020 questionnaire.
\*\*This question included the UK government's provisional prioritisation list for Covid-19 vaccination for participants to review.
\*\*\*This question was deliberately posed as the last item in the questionnaire as not to bias the free-text responses described in table 2.

In response to the question of which groups should be prioritised, unrestricted free-text responses (7,838) indicated that teachers (988, 12.6%), Black, Asian and Minority Ethnic (BAME) groups (837, 10.7%), general key workers (807, 10.3%) children (582, 7.4%), and university students (529, 6.7%) were the most cited. 32.6% were concerned that the priority list made no reference to BAME groups (table 2).

**Table 2.**
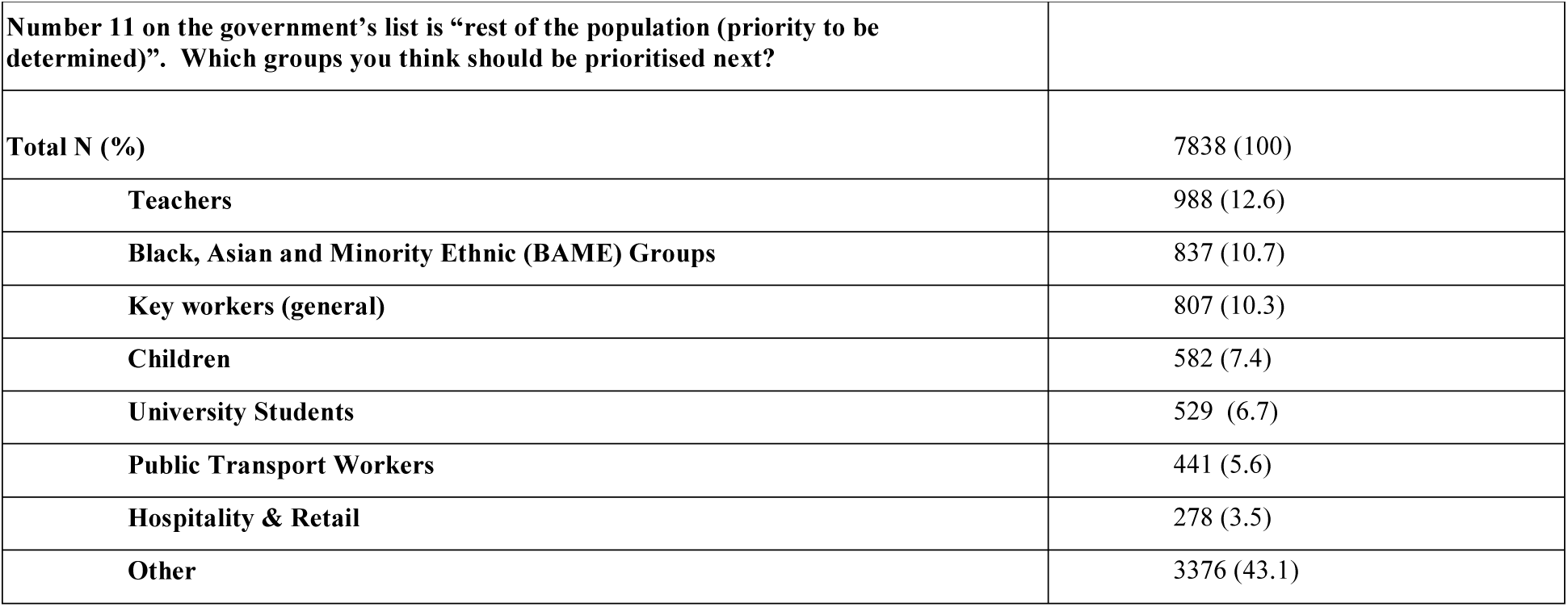
Free-text analysis of next-in-line prioritisation.

### Second, Follow-up Questionnaire (December 31st, 2020)

Among 9617 responses (51.8 % response rate, including 78.6% of participants from the first questionnaire), baseline acceptance of vaccination was 81.2%, increasing to 85.1% with the addition of 375 participants who changed their minds to wanting vaccination in light of news of the new variant. 1203 (12.5%) would want to significantly change their behaviour within a few weeks of completing vaccination, compared to 2077 (21.6%) who would want no change even if cases were falling (1864, 19.4%) or at very low levels (4317, 44.9%) (table 3).

**Table 3.**
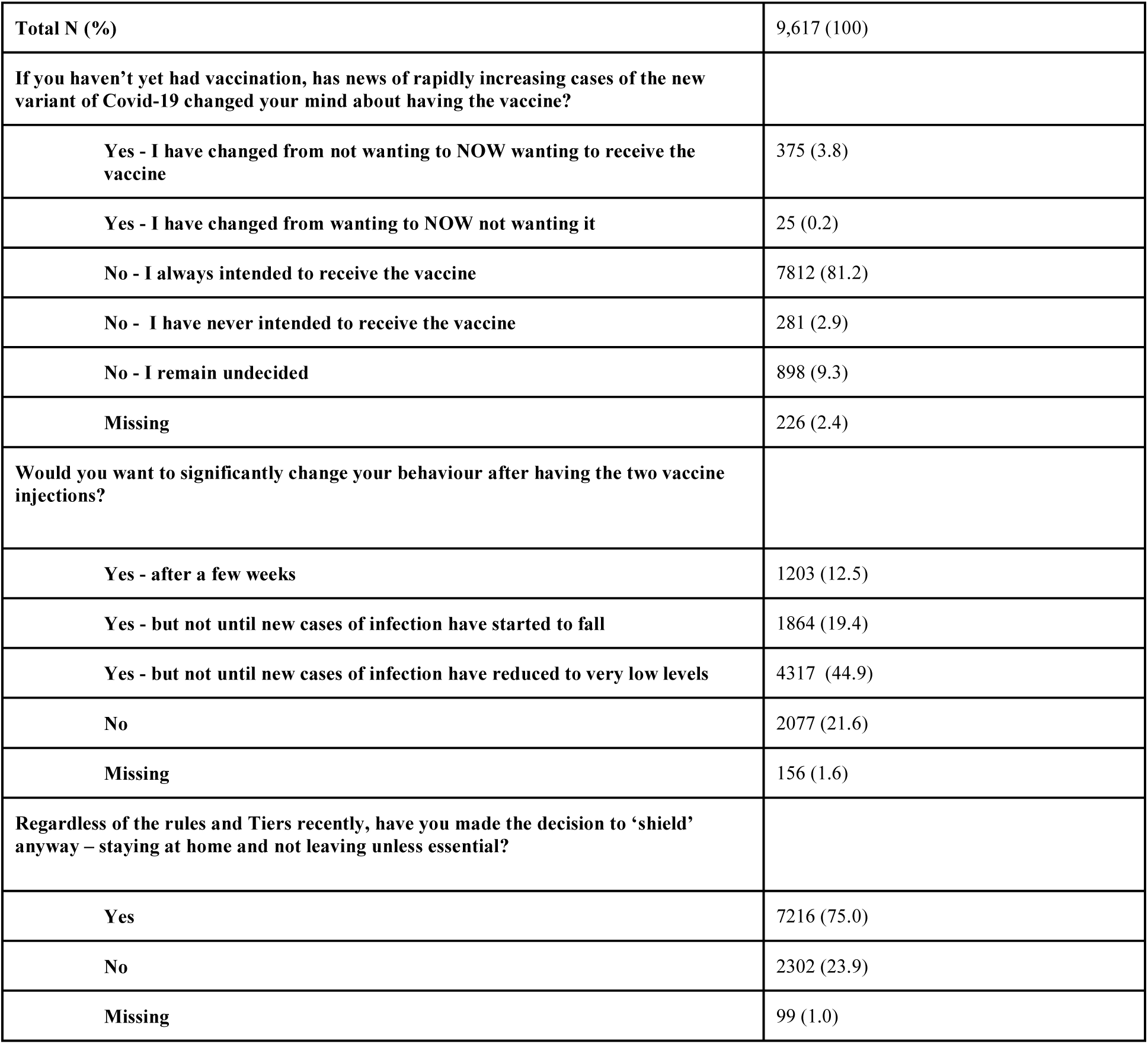
Second, Follow-up Questionnaire (December 31st 2020)

|  |  |
| --- | --- |
| **Total N (%)** | 9,617 (100) |
| **If you haven't yet had vaccination, has news of rapidly increasing cases of the new variant of Covid-19 changed your mind about having the vaccine?** |  |
| Yes - I have changed from not wanting to NOW wanting to receive the vaccine | 375 (3.8) |
| Yes - I have changed from wanting to NOW not wanting it | 25 (0.2) |
| No - I always intended to receive the vaccine | 7812 (81.2) |
| No - I have never intended to receive the vaccine | 281 (2.9) |
| No - I remain undecided | 898 (9.3) |
| Missing | 226 (2.4) |
| **Would you want to significantly change your behaviour after having the two vaccine injections?** |  |
| Yes - after a few weeks | 1203 (12.5) |
| Yes - but not until new cases of infection have started to fall | 1864 (19.4) |
| Yes - but not until new cases of infection have reduced to very low levels | 4317 (44.9) |
| No | 2077 (21.6) |
| Missing | 156 (1.6) |
| **Regardless of the rules and Tiers recently, have you made the decision to 'shield' anyway – staying at home and not leaving unless essential?** |  |
| Yes | 7216 (75.0) |
| No | 2302 (23.9) |
| Missing | 99 (1.0) |

## DISCUSSION

In this large, UK-wide sample of NHS patients, there was a 15% increase in acceptance of Covid-19 vaccination in the critical 50 days of case escalation leading to the UK Government’s decision for the New-Year lockdown compared with the first week reporting an effective vaccine. The increased intent to receive the vaccine from 70 to 85% was attributable in one third to the impact of the new variant, with 75% of respondents “shielding” – staying at home and not leaving unless essential – regardless of health status or tier rules. The growing impact of personal choice among the increasingly informed public also revealed that 12.5% of respondents intend to lift restrictions on their lives two weeks after completing vaccination, compared with 45% intending to do so only when cases have reduced to a low level.

Although the duration of individual protection from the UK’s expanding Covid-19 vaccination programme remains unknown, assuming this to be one year, previous modelling by Anderson et al.^3^ indicates critically that the increase to 85% remains inadequate for herd immunity alone to prevent Covid-19 remaining endemic. We therefore identify factors to inform the public health messaging to improve uptake: younger people and females, in combination most exposed to Covid-19^9^, should be proactively targeted.

That 33% of respondents were concerned that the priority list makes no reference to BAME groups presents a particular challenge to the UK government, given evidence that this group are not only at increased risk from Covid-19 infection, but also have historically had lower uptake of existing vaccination programmes.^10,11^ Our results indicate not only the priority considerations for public health messaging necessary to increase the inadequate level of intended uptake, but also the public’s concern at omission of reference to BAME groups and a preference for extending the prioritisation list in the first instance to teachers and BAME groups. Listening to these preferences as part of a wider public health strategy will help build trust and community engagement, critical if achieving herd immunity as quickly as possible is to become a realistic possibility and to encourage compliance with other public health measures.

The limitations of this study include that although the LoC-19 cohort of NHS patients engaging with questionnaires is large, it is unlikely to be representative of the whole UK population, limiting the generalisability of our findings. Nonetheless, smaller studies have also reported between 65-85%^12,13^ Covid-19 vaccine acceptance rates. Importantly, our sample is unique in comprising NHS patients and therefore incorporating a spectrum of individuals most at risk from Covid-19 and therefore a demographic mix of the most important group to study for informing Covid-19 vaccine health policy.

## Conclusion

In this population, Covid-19 vaccine hesitancy has decreased over time, partly due to news of a more infectious new variant, but vaccine acceptance is still below levels that would enable progress towards herd immunity. Participatory community engagement should be part of a strategy to improve uptake by considering the public’s preferences, such as those expressed here that teachers and BAME groups should be prioritised.

**What is already known on this topic**

- Effective public health messaging can mitigate Covid-19 vaccine hesitancy, the critical modifiable variable for progressing towards herd immunity
- Since news of the the first efficacious vaccine, vaccination programmes have commenced against an uncertain backdrop of new variants and unclear prioritisation plans beyond the immediately highest-risk groups

**What this study adds**

- This longitudinally participatory epidemiology study of >9,000 NHS patients informs Covid-19 vaccine hesitancy and behaviour change across a critical 50 day period of the pandemic
- The emergence of new Covid-19 variants motivated a decline in vaccine hesitancy, but with overall uptake likely still below levels to advance herd immunity; we also highlight preferences for prioritising teachers and BAME groups

## Data Availability

Data availability: Imperial College Healthcare NHS Trust is the data controller. The datasets analysed in this study are not publicly available but can be shared for scientific collaboration subject to meeting requirements of the institution's data protection policy.

## Author Contribution

Patrik Bachtiger: study design, data collection, literature review, data analysis, figures, writing Alexander Adamson: study design, literature review, figures, data analysis, writing

William A Maclean: figures, data analysis, writing

Jennifer K Quint: study design, literature review, data analysis, figures, writing

Nicholas S Peters: study design, data collection, literature review, data analysis, figures, writing

NSP is the guarantor. The corresponding author attests that all listed authors meet authorship criteria and that no others meeting the criteria have been omitted.

The lead author affirms that this manuscript is an honest, accurate, and transparent account of the study being reported; that no important aspects of the study have been omitted; and that any discrepancies from the study as planned (and, if relevant, registered) have been explained.

## Data availability

Imperial College Healthcare NHS Trust is the data controller. The datasets analysed in this study are not publicly available but can be shared for scientific collaboration subject to meeting requirements of the institution’s data protection policy.

## Ethical approval

The weekly questionnaire was a direct care tool for patients to self-monitor their wellbeing during the Covid-19 pandemic. Review by the Imperial College Healthcare NHS Trust Data Protection Office advised ethical approval for data analysis and publication was not required. Participants were informed prior to completing responses that these would be anonymised to inform local and national health policy and were free to opt out.

## Patient and Public Involvement

Study participants (patients) were involved in the review of questionnaire items before being finalised for inclusion.

