## supplementary for "Increasing but inadequate intention to receive Covid-19 vaccination over the first 50 days of impact of the more infectious variant and roll-out of vaccination in UK: indicators for public health messaging"

**SUPPLEMENTARY MATERIALS**

**Supplementary Figure 1.** Map of UK showing CIE registrants by postcode (each green dot indicating at least one registrant in that postcode area).

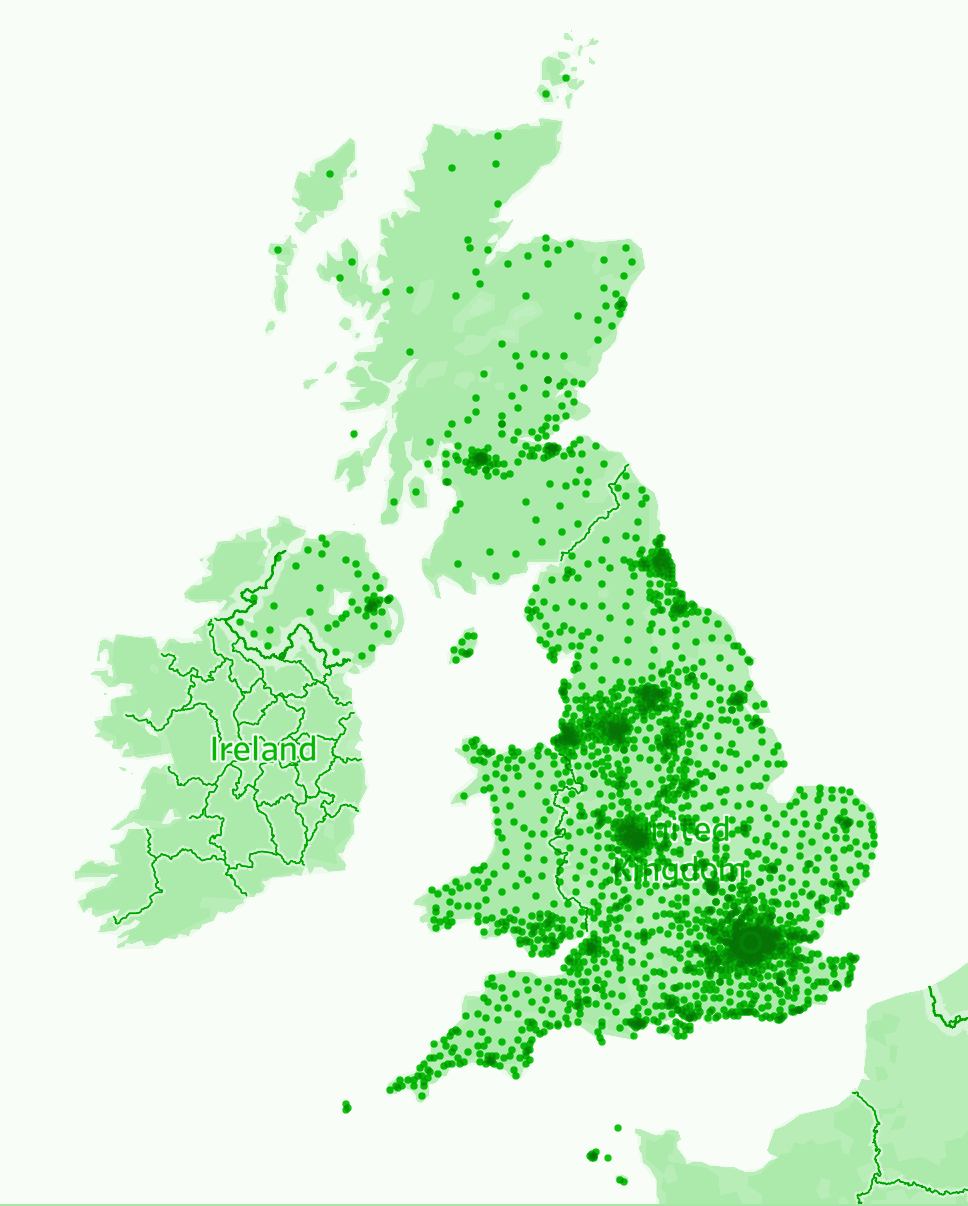

**Supplementary Table 1.** UK Government Covid-19 vaccination priority list, as presented to participants.

The government has issued the following provisional ranking of priority for a COVID-19 vaccine:

1. Older adults’ resident in a care home and care home workers
2. All those 80 years of age and over, and health care workers and social care workers
3. All those 75 years of age and over
4. All those 70 years of age and over
5. All those 65 years of age and over
6. High-risk adults under 65 years of age
7. Moderate-risk adults under 65 years of age
8. All those 60 years of age and over
9. All those 55 years of age and over
10. All those 50 years of age and over
11. Rest of the population (priority to be determined)

Source: Joint Committee on Vaccination and Immunisation. Advice on priority groups for COVID-19 vaccination, 30 December 2020. <https://www.gov.uk/government/publications/priority-groups-for-coronavirus-covid-19-vaccination-advice-from-the-jcvi-2-december-2020/priority-groups-for-coronavirus-covid-19-vaccination-advice-from-the-jcvi-2-december-2020>
